# IS SHORT-TERM ACYCLOVIR TREATMENT RELATED WITH MORE UNFAVOURABLE OUTCOMES IN PATIENTS WITH HSV-1 ENCEPHALITIS?

**DOI:** 10.1101/2022.02.08.22270584

**Authors:** Ferhat Arslan, Ravza Akgündüz, Abdülkadir Ermiş, Begüm Bektaş, Ebru Erbayat, Haluk Vahaboğlu

## Abstract

**Background:** Herpes simplex encephalitis (HSE) is the most common form of sporadic encephalitis which is caused by herpes simplex virus type 1 (HSV-1). Current guidelines recommend intravenous Acyclovir for 14–21 days in cases of HSE.

**Objectives:** Optimizing the Acyclovir treatment duration is important in regards to preventing Acyclovir-related neurotoxicity and nephrotoxicity.

**Study design:** We retrospectively evaluated 13 patients, who were diagnosed with HSE by molecular testing (HSV-1 PCR positivity in cerebrospinal fluid), in two university hospitals in Istanbul, between 2010 and 2021. The patients were treated either 10 days or less as a short-term treatment regimen of Acyclovir or for 14 days or more as long-term treatment regimen

**Results:** The median age was 58 years (range 24–82 years) and 54% of them were male. The median follow-up time was 79 days (range 20-670 days) after discharge. Long-term treatment was used in 6 and a short-term treatment regimen was used in 7 cases. One of the patients died on the 4th day of Acyclovir treatment. One patient never received Acyclovir treatment. Of the 5 patients who received long-term treatment, 3 (21-28 days) had amnesia and orientation-cooperation restriction, one (21 days) died on the 2nd day of treatment, while the other (14 days) had no sequelae. There were no sequelae in three out of 5 patients who received short-term treatment.

**Conclusions:** We believe that it is necessary to determine the optimal Acyclovir therapy duration should be evaluated in a prospective randomized clinical trial in large patient population.

## INTRODUCTION

Herpes simplex encephalitis (HSE) is the most common form of sporadic encephalitis which is caused by herpes simplex virus type 1 (HSV-1). HSV-1 generally infects orolabial mucosa and face skin epithelium, gains access to neuronal axonal ending, and then is transported to the trigeminal ganglia (TG)^1^. It is estimated approximately 200 million recurrent HSV-1 related infections develop every year^2^. However, very few of these cases are HSE. The rareness might be due to its different ways of inoculation in the brain and TG or due to the primary infection rather than reactivation^3^. Despite the increasing knowledge, there are still unclear points in the pathogenesis of HSE.

Although it was reported that the mortality rates were above 70% and the rates of neurological sequelae were around 90%, it has now been decreased to 30% for both mortality and severe neurological sequelae^4^. Mortality rates have decreased significantly by the discovery of Acyclovir, improvements in radiological screening (cranial magnetic resonance imaging), and the use of molecular (HSV-1 DNA) diagnostic methods in medical practice. Vidarabine was used as a comparative drug in two double-blind, randomized controlled clinical trials investigating the efficacy and side effects of Acyclovir treatment for HSE^5,6^.

The duration of Acyclovir treatment was ten days in these trials. However, current guidelines recommend intravenous Acyclovir for 14–21 days in cases of HSE^7,8^. Optimizing the Acyclovir treatment duration is important in regards to preventing Acyclovir-related neurotoxicity and nephrotoxicity ^9^. Here, we aimed to discuss the recommended treatment durations in patients with HSE by using short-term versus long-term acyclovir treatment. We also emphasize the effects of adjunctive therapies, such as steroids and antibiotics, on these cases.

## MATERIALS AND METHODS

### Patients

We retrospectively evaluated 13 patients, who were diagnosed with HSE by molecular testing (HSV-1 PCR positivity in cerebrospinal fluid), in two university hospitals in Istanbul, between 2010 and 2021. Their clinical (altered mental status, seizures or focal neurological symptoms) and radiological findings (detection of lesions in diffusion or contrast-enhanced MRI), were analyzed. The patients were treated either 10 days or less as a short-term treatment regimen of Acyclovir or for 14 days or more as long-term treatment regimen. Chronic Kidney Disease Epidemiology Collaboration (CKD-EPI) formula was used for the assessment of the renal damage for each case.

Ethics committee approval was obtained from the ethics committee of Istanbul Medeniyet University.

The neurological status for each case was noted from the outpatient follow-up records. Neurological sequelae have been defined as any damage to the central nervous system that results in cognitive, sensory, or motor deficits, which may also manifest as long-term emotional instability and seizures in severe cases^10^.

## RESULTS

### Clinical and MRI features

A summary of the clinical information of 13 patients is shown in **Table 1**.

**TABLE 1:** CLINICAL FEATURES OF PATIENTS WITH HERPES SIMPLEX ENCEPHALITIS (HSE) WHO RECIEVED ACYCLOVIR TREATMENT

|  | Age range /Sex | Symptoms | Signs | Cranial MRI findings | CSF PCR | Time to Treatment<sup>a</sup> | Outcome |
| --- | --- | --- | --- | --- | --- | --- | --- |
| **Patient 1** | (50-55)/F | Altered mental status, dysarthria, behavioral changes | Confused, disoriented, limited cooperation | Bilateral insular cortex, right temporal lobe, contrast enhancement and diffusion restriction | CSF HSV Type-1 PCR: Positive | 1 day | No sequelae alive |
| **Patient 2** | (80-85)/M | Fever, headache | Lethargy | Right temporal lobe, hippocampal, insular cortex contrast enhancement, and diffusion restriction | CSF HSV Type-1 PCR: Positive | None | No sequelae alive |
| **Patient 3** | (75-80)/F | Altered mental status, dysarthria, difficulties on walking | Unconscious GCS=10 | Bilateral temporal lobe (right dominant), contrast enhancement and diffusion restriction | CSF HSV Type-1 PCR: Positive | 3 days | No sequelae alive |
| **Patient 4** | (40-45)/M | Altered mental status, dysarthria, fever, headache, nausea and vomiting, loss of balance | Confused, disoriented, limited cooperation | Right temporal lobe, parasylvian, hippocampal and parahippocampal contrast enhancement and diffusion restriction | CSF HSV Type-1 PCR: Positive | 7 days | No sequelae alive |
| **Patient 5** | (65-70)/F | Fever, Headache, Dysarthria | Confused, disoriented, limited cooperation | Left temporal lobe, hippocampal, insular cortex contrast enhancement and diffusion restriction | CSF HSV Type-1 PCR: Positive | 5 days | No sequelae alive |
| **Patient 6** | (20-25)/F | Fever, headache, altered mental status, nausea and vomiting | Confused, disoriented, limited cooperation | Left temporal lobe, left insular cortex, uncut and hippocampus gyrus, dorsal thalamus contrast enhancement and diffusion restriction | CSF HSV Type-1 PCR: Positive | 3 days | Loss of balance, seizures alive |
| **Patient 7** | (65-70)/M | Altered mental status, convulsion | Confused, disoriented, limited cooperation | Right temporal lobe, insular cortex T2 hyperintensity and diffusion restriction | CSF HSV Type-1 PCR: Positive | 5 days | Dead |
| **Patient 8** | (20-25)/F | Fever, Headache, Seizure | Lethargy | Right temporal lobe, insular cortex, thalamus contrast enhancement and diffusion restriction | CSF HSV Type-1 PCR: Positive | 3 days | No sequelae alive |
| **Patient 9** | (70-75)/M | Altered mental status, headache, vesicular rash | Lethargy | Normal | CSF HSV Type-1 PCR: Positive | 10 days | Dead |
| **Patient 10** | (40-45)/M | Altered mental status, Fever, Decreased hearing, | Confused, disoriented, Lethargy diplopia | Right Frontotemporal lobe, T2 hyperintensity | CSF HSV Type-1 PCR: Positive | 7 days | Dead |
| **Patient 11** | (60-65)/F | Altered mental status, Convulsion, Fever, Headache | Confused, disoriented, limited cooperation | Left temporal lobe, insular cortex, contrast enhancement and diffusion restriction | CSF HSV Type-1 PCR: Positive | 8 days | Confused, loss of time orientation, alive |
| **Patient 12** | (65-70)/M | Altered mental status,<br>Fever,<br>Convulsion | Confused,<br>disoriented,<br>limited cooperation | Bilateral temporal lobe (right dominant),<br>Right thalamus contrast enhancement and diffusion restriction | CSF HSV Type-1 PCR: Positive | 4 days | Confused, prolonged reaction time to complex orders, alive |
| **Patient 13** | (60-65)/M | Altered mental status,<br>Headache,<br>behavioral changes | Confused,<br>disoriented,<br>limited cooperation<br>sensory aphasia | Right Fronto-temporal lobe, contrast enhancement and diffusion restriction | CSF HSV Type-1 PCR: Positive | 8 days | Confused, disoriented, alive |

<sup>a</sup>: Time from symptoms onset to treatment initiation

The median age was 58 years (range 24–82 years) and 54% of them were male. The median follow-up time was 79 days (range 20-670 days) after discharge. Five patients had an underlying disease-causing immune deficiency: Two had organ tumors (Lung cancer and prostate cancer, other two patients had Diabetes mellitus, which one of them with febrile neutropenia due to hemophagocytic syndrome and one patient had recurrent orolabial Herpes reactivation at the time of the diagnosis. The median duration of diagnosis, starting from the symptom onset was 5 days (IQR 5 days Q3:8 Q1:3 IQR:5). The CSF analysis showed lymphocytic pleocytosis in 9/13 patients (median leukocyte count 161 cells/mL, range 0– 500), and elevated protein concentration in 10/13 (median 91 mg/dL, range 35–349) While unconsciousness was observed in all of the patients, no neck stiffness was found in any of them. On admission, fever (9/13), headache (9/13), seizures (7/13) and focal neurological signs (diplopia and hearing loss in 1 patient and right hemiparesis in another patient) were noted.

In all patients, the clinical diagnosis of HSE was confirmed by the MRI findings of unilateral (9 patients) or bilateral (3 patients) increased fluid-attenuated inversion recovery (FLAIR)/T2 signal abnormalities in temporal regions. In five patients, frontal (2 patients), hippocampal (2 patients) and occipital (1 patient) FLAIR/T2 signal abnormalities were additionally observed.

### Treatment

Long-term treatment was used in 6 and a short-term treatment regimen was used in 7 cases. One of the patients died on the 4th day of Acyclovir treatment. One patient never received Acyclovir treatment. Twelve patients had also broad-spectrum antibiotic treatment at the beginning. Adjunctive steroid therapy was given in 3 patients. Antiepileptic therapy was needed for 10 patients. Treatment modalities and follow-up of renal functions are summarized in Table 2

**TABLE 2:** TREATMENT MODALITIES AND RENAL FUNCTIONS OF PATIENTS WITH HERPES SIMPLEX ENCEPHALITIS (HSE)

| **Patients** | **Empirical antibiotics** | **Acyclovir start time <sup>a</sup>** | **Acyclovir duration** | **Steroid treatment** | **Antiepileptic treatment** | **CNS Symptoms resolved time** | **eGFR(CKD-EPI) on admission** | **eGFR(CKD-EPI) on discharge** |
| --- | --- | --- | --- | --- | --- | --- | --- | --- |
| Patient 1 | Cefuroxime axetil,<br>Meropenem-Linezolid-INH-Rifampin | 1 d | 10 d | 2 d | 1 months after discharge | 3 days (AAS) | 97 | 86 |
| Patient 2 | Meropenem<br>Linezolid | None | None | None | None | 14th days of admission | 45 | 76 |
| Patient 3 | Ampicillin | 3 d | 9 d | None | 5 months after discharge | 9 days (AAS) | 36 | 55 |
| Patient 4 | Ceftriaxone | 1 d | 7 d | None | 2 months after discharge | 4 days (AAS) | 117 | 68 |
| Patient 5 | Amoxicilline-Clavulanate,<br>Ampicillin-Ceftriaxone | 5 d | 7 d | None | None | 4 days (AAS) | 83 | 55 |
| Patient 6 | Ceftriaxone<br>Ampicilline | 1 d | 10 d | 7 d + 20 d | 2nd day of<br>hospitalization | 3 days (ASI) | 127 | 134 |
| Patient 7 | Piperacilline-Tazobactam | 5 d | 4 d | None | 2nd day of<br>symptoms | None | 100 | 112 |
| Patient 8 | Ceftriaxone | 1 d | 14 d | 3 d | 2nd day of<br>hospitalization | 10th days of<br>admission | 131 | 123 |
| Patient 9 | Ceftriaxone | 1 d | 21 d | None | None | 12th days of<br>admission | 85 | 82 |
| Patient 10 | meropenem | 7 d | 21 d | 10 d | None | 14 days (AAS) | 70 | 36 |
| Patient 11 | Ceftriaxone | 8 d | 28 d | None | 1st day of<br>hospitalization | 5 days (AAS),<br>but<br>confusion<br>perminent | 74 | 95 |
| Patient 12 | Ceftriaxone and<br>levofloxacin | 4 d | 24 d | None | 1st day of<br>hospitalization | 7 days (AAS)<br>but<br>confusion<br>perminent | 81 | 102 |
| Patient 13 | None | 8 d | 21 d | None | 3rd day of<br>hospitalization | 3 days (AAS) | 72 | 62 |

<sup>a</sup>: Time from symptoms onset to treatment initiation
CNS: Central nervous system; AAS: After acyclovir stopped; ASI: After steroid initiation

### Outcomes

Death occurred in 2 patients. One of them was already on severe immunosuppressive therapy due to EBV-related hemophagocytosis. The other patient had lung cancer with brain metastases and subarachnoid hemorrhage. Of the 5 patients who received long-term treatment, 3 (21-28 days) had amnesia and orientation-cooperation restriction, one (21 days) died on the 2nd day of treatment, while the other (14 days) had no sequelae. There were no sequelae in three out of 5 patients who received short-term treatment, however, balance problems continued in one of them in 6 months after treatment. Neurological sequelae did not develop in the patient who did not receive any treatment. CKD-EPI decrease associated with renal failure was seen only in one patient.

## DISCUSSION

There is a solid consensus for the early initiation of effective antiviral agents in HSE, however, recommendations regarding the duration of the antiviral treatment is still at the level of expert opinion. Although this is a case series study and it is consisted of a small number of cases, we observed some interesting results: In short term Acyclovir treatment group, CNS symptoms of all 6 patients improve within the first week of the drug discontinuation, contrary to the long-term treatment group, where the symptoms and signs of the HSE continued after the discontinuation. In another word, neurological sequelae in long-term treatment group were more common than the short-term treatment group. Additionally, the patients who received steroid treatments from both groups (patient 1,6,7,10) did not reveal any neurological deterioration. Seizure in one patient in 26th month and progressive amnesia in another patient on 5th month of the follow-up after short term treatment, respectively. Interestingly, neurological sequelae did not develop in the patient (patient 2 who was not given acyclovir treatment. One out of patients were treated with empirical antibiotics. Our observations may encourage steroid and short-term Acyclovir therapy in the management of the patients with HSE.

Stevens et al. showed that HSV viral replication continued for up to 8 days in a spinal ganglion of animal model study^11^. In a superior cervical ganglia (*SCG*) cell cultures-based HSV-1 model study, it was shown that phase 1 and phase 2 transcriptional processes take place in less than 5 days^12^. Toll like receptor 3 (TLR-3) dependent production of IFN-α/β and IFN-λ is seen as a protective factor for HSE in children^13^. Additionally, Acyclovir doesn’t have an inhibitor effect on the replication of the HSV-1 in human neural cell lines without human interferon-alpha (IFN-alpha)^14^. IFN-β treatment in baby mice with HSV encephalitis model decreased neuro-invasion and increase the survival rate^15^. We may conclude that the combined effect of Acyclovir implementation and host interferon response can be achieved only at the early days of the HSE. In our study, the median time from symptoms onset to treatment initiation was 3.4 days in the group receiving short-term acyclovir treatment, while it is 6.6 days in the group receiving long-term treatment. This may contribute to the low rate of neurological sequelae in the group receiving short-term treatment.

Other probable justifications have been put forward for long-term antiviral treatment rationale. In situ low-level HSV-1 replication hypothesis examined in the prospective randomized double-blind study of Gnann et al, long-term Valacyclovir treatment was found to have no significant effect on neurological outcome^16^. The second issue is the assumed virological relapse that may be seen after the early cessation of acyclovir. In a long-term study of 32 adult HSE cases prospectively observed, HSE relapse was reported in 4 patients. However, HSV PCR positivity was not detected in any of the 12 CSF samples taken from these patients. It has been reported that putative relapses occur within the first 4 months after infection^17^. It was reported that HSV-1 antigen and HSV-1 DNA were detected immunohistochemically in the necrotic right frontal and parietal lobe samples from the autopsy of a patient who presented with status epilepticus 5 years after HSE^18^. The authors claim that this case is the first pathologically and molecularly confirmed relapse of HSE in the medical literature. However, progressive neurological deterioration throughout 5 years in the course of the case raises questions about whether the patient is in a relapse or a chronic autoimmune process^19,20^.

During the SARS-CoV-2 pandemic, there has been increased awareness about the possible antiviral effects of some drugs that are not used as virucidal^21^. Empirical antibiotics and steroids along with the Acyclovir treatment is a quite common clinical approach to the HSE cases in the first days before definitive diagnosis^22^. Inhibitory activity of Cephalosporin derivatives on HSV-1 was shown in vitro^23^. Similarly, some other molecules (Valproic acid, Valpromide and Valnoctamide) that inhibit the replication of HSV-1 virus in oligodendrocytes have been identified^24^. The researchers emphasized that these molecules could be an effective and safe treatment alternatives in HSE. The effect of non-antiviral drugs, especially antibiotics on HSE prognosis has not been studied.

Researchers have developed human induced pluripotent stem cells (hiPSCs) and neuronal cells based on 3D organoid models to establish HSV-1 infections^25^. Although in vitro establishment of neuron cells infection has been shown, there are in vivo differences. HSV-1 has evolved latency mechanisms to stay in neuron cells in order to escape from immunity^26^. Due to the logic of co-evolution, the relationship between HSV and neuronal cells cannot be considered to be direct damage (cytopathic damage). The important question here is which cells are susceptible for HSV-1 replication in brain^27^. As we understand from the CSF finding, erythrocytosis, the blood-brain barrier (BBB) is disrupted in HSE patients^28^. Brain microvascular endothelial cells may play the primary role in neuronal damage due to HSV-1 induced endothelial cell adhesion molecules that interact with lymphocytes and neutrophils recruitment^29,30^. We may speculate that steroids may play a role to diminish this recruitment and decrease the vascular inflammation as adjunctive therapy. Adjunctive steroid therapy initiation timing and its duration are still elusive^31^.

In conclusion, the optimal duration of antiviral treatment for HSE is still a controversial issue. Those who received short-term treatment might not have worse outcome than those who received long-term treatment in terms of neurological sequelae. We believe that it is necessary to determine the optimal Acyclovir therapy duration, as well as the steroids’ effect on probable vasculitis and should be tested in a prospective randomized clinical trial in large patient population.

## Data Availability

All data produced in the present study are available upon reasonable request to the authors

